# Respiratory syncytial virus (RSV) notifications and trends in the transmission cycles from infants and young children to older adults in Ireland: an analysis of incidence shifts over a decade

**DOI:** 10.64898/2025.12.01.25341394

**Authors:** Roy K Philip, Kaushik Mangroo, Natalie Gendy, Helen Purtill, Eva Kelly, Lisa Domegan, Maureen O’Leary

**Author notes:** Corresponding Author: Prof Roy K Philip, Adjunct Full Professor of Neonatology, University Maternity Hospital Limerick (UMHL) and University of Limerick School of Medicine, Limerick, V94 C566, Ireland.

## Abstract

**Objectives:** Understanding the epidemiological shifts of respiratory syncytial virus (RSV) is essential to inform public health interventions, particularly given its increased burden on healthcare systems post-COVID-19 pandemic. This study aimed to examine age-specific trends and seasonal variations in RSV incidence, considering the recent introduction of a newborn RSV immunisation programme in Ireland.

**Design:** A surveillance time series study analysing routinely collected RSV notification data.

**Settings:** National-level weekly RSV notifications collected by the Health Service Executive (HSE)-Health Protection Surveillance Centre (HPSC) in Ireland from 2012 to 2024.

**Participants:** Infants (<1 year), young children (1-4 years), and older adults (≥65 years) with laboratory-confirmed RSV, from within the corresponding Irish population.

**Outcome measures:** Annual trends in RSV epidemiology with special reference to the pre- and post-COVID-19 winter surges, and the time lag in age-related transmission to peak incidence among the various age groups. Data were analysed to evaluate incidence rates, peak timing, age-related transmission trends, and lag times before and after the COVID-19 pandemic.

**Results:** The study examined the increasing incidence of RSV post-COVID and a significant shift toward earlier RSV peaks in recent years (2021/2022, 2022/2023, and 2023/2024 seasons) in Ireland, with the onset and peak of the season nearly two months earlier than in pre-COVID pandemic seasons (p < 0.01). Cross-correlation factor (CCF) analysis indicated a sequential spread of RSV infections, where a peak in older adults followed an initial rise in cases among infants and young children, within a 3-5 week period (maximum cross-correlation = 0.86 at lag 4 weeks, p<0.001). Post-pandemic, infants exhibited higher infection rates, with incident rates significantly higher in all seasons post-COVID (p<0.001) and peak intensities increasing by over 60% from 2021/22 to 2023/24.

**Conclusion:** This analysis highlights an early seasonal onset and intensified RSV burden among infants in recent winters (2021/2022, 2022/2023, and 2023/2024 seasons). Quantifying the time lag for the community-level RSV transmission from infants and young children to older adults will offer insights to optimise RSV intervention strategies as a *‘life-course approach’* to alleviate healthcare system pressures during peak seasons.

**Data Availability Statement:** All data relevant to the study are included in the article or uploaded as supplementary information. Weekly RSV notification data reported by the HSE-Health Protection Surveillance Centre (HPSC) from week 40, 2012 to week 20, 2024 is available at https://respiratoryvirus.hpsc.ie/

**Strengths and limitations of this study:**

- A national, prospectively collected weekly surveillance dataset spanning twelve seasons (pre- and post-COVID-19) provided a robust basis for analysing RSV trends in Ireland.
- Age-stratified analysis (<1 year, 1–4 years, ≥65 years) allowed comparison of temporal patterns across key population groups.
- Application of time-lag correlation methods quantified inter-age transmission lags, supporting community-level planning and preparedness.
- The study establishes a national pre-immunisation epidemiological baseline to inform assessment of future RSV immunisation programmes.
- The absence of national data on RSV-related general practitioner (GP) visits, hospitalisations, or critical-care demand limited evaluation of the full clinical burden.

## INTRODUCTION

Respiratory Syncytial Virus (RSV) is a leading cause of severe acute lower respiratory infections (ALRI), placing a significant burden on healthcare systems worldwide and driving high rates of morbidity, hospitalisations, and mortality, especially among children under five years and older adults over 65 years.^1^ ^2^ ^3^ ^4^ ^5^ Globally, RSV accounts for approximately 33 million cases, 3.6 million hospital admissions, and nearly 26,300 in-hospital deaths annually among young children.^6^ RSV is the leading cause of hospitalisation in infancy, and nearly two-thirds of infections occur during the first four months of life.^7^ Although prematurity and congenital heart disease are well-established risk factors for severe infections, most of the healthcare burden arises from infections in otherwise healthy term-born infants under six months, with perinatal and sociodemographic factors also playing a role in RSV-associated ALRI.^1^ ^5^ ^8^ ^9^ Lack of breastfeeding, overcrowding and exposure to cigarette smoke increase the incidence and severity of RSV-associated ALRI.^4^ ^10^ ^11^ ^12^

In older adults, the burden is equally concerning, with 10-31% of hospitalised RSV cases requiring intensive care and 3-17% needing mechanical ventilation, especially among high-risk groups with comorbidities such as cardiopulmonary disease.^7^ However, accurately assessing RSV-related morbidity and mortality in this population remains challenging due to comorbidities, atypical symptoms, and lack of uniform viral testing which affects diagnosis and severely underestimates its true burden on this vulnerable population, compared to the paediatric population.^.5 6^ In older adults in Europe, the morbidity burden of RSV may be comparable to that of influenza and presents with significant clinical severity and complications.^6^ ^7^

In temperate climates, RSV follows predictable seasonal patterns, peaking in winter months in the Northern Hemisphere and mid-year in the Southern Hemisphere.^5^ ^8^ ^13^ Despite links between RSV seasonality and climatic factors, such as temperature and humidity, a comprehensive understanding of these patterns remains elusive.^13^ ^14^ 15

During the COVID-19 pandemic, stringent public health measures, including periods of ‘lockdown’ led to a 98% reduction in RSV cases during 2020-2021.^5^ ^16^ ^17^ ^18^ However, the post-pandemic period, starting with the 2021-2022 season has been marked by a stark resurgence in RSV infections, with peaks occurring earlier than expected and viral transmission extending throughout the summer months of 2021.^5^ ^12^ ^19^ ^20^ The increased RSV incidence rates observed in 2021/2022, 2022/2023 and 2023/2024 seasons were multifactorial and likely due to *‘immunity debts’* often resulting in a ‘*tripledemic*’ or ‘*tridemic*’ (concurrent peaking of RSV, influenza and SARS-CoV-2) have increased significant pressures on healthcare systems.^5^ ^9^ ^21^ ^22^ Increased use of multiplex PCR testing and laboratory capacity since the COVID-19 pandemic must also be considered when comparing respiratory virus surveillance data pre- and post the COVID-19 pandemic.

Given the rising RSV burden and the recent approval by European Medicines Agency (EMA) of various RSV immunisation with monoclonal antibodies and RSV vaccines, there is an urgent need for effective prevention strategies that align with RSV’s epidemiological trends. At the recommendation of the National Immunisation Advisory Committee (NIAC) the Department of Health requested the Health Service Executive (HSE) to initiate the RSV immunisation programme for all newborn infants in Ireland.^23^ This immunisation approach designated as a ‘*Pathfinder Programme*’ encompasses intramuscular administration of nirsevimab, a long-acting monoclonal antibody (mAb) to all newborn infants born between 1^st^ September 2024 to 28^th^ February 2025, aimed at reducing the RSV-associated hospital admissions and critical care (paediatric high dependency unit-PHDU and paediatric intensive care unit-PICU) demand for infants.^23^ ^24^ ^25^ This approach aligns with our 12-year study of RSV epidemiology across infants and older adults in Ireland, offering an opportunity to examine transmission dynamics across age groups, where RSV spreads from younger to older populations, often within households, a phenomenon known as *‘RSV march’*.^26^ ^27^ ^28^ ^29^ As Ireland considers long-term strategies for RSV immunisation, understanding RSV epidemiology, transmission trends and disease burden in vulnerable groups is vital for effective immunisation strategies.

The circulation of RSV in the community and the resultant illness burden among the high-risk age groups (<1 year, 1-4 years, and ≥65 years) also resonates with the priority of the World Health Organisation (WHO) on a life-course immunisation (LCI) approach.^30^ With the older adult population for the first time in history surpassing <5-year-old population globally, a comprehensive life-course approach in population health is warranted more than ever before.^31^ A cohesive epidemiological approach to quantify the time lag in RSV seasonality and peak incidence from 0-4 years to ≥65 years would be valuable for modelling, predicting, and planning for the winter surges that result in increased emergency department (ED) visits and demand on the limited acute bed stock during the winter months.^.32^

This paper investigates the national RSV surveillance notifications in Ireland from week 40 in 2012 to week 20 in 2024, focusing on incidence trends across age groups: infants (<1 year), young children (1-4 years), and older adults (≥65 years). By examining seasonal shifts before, during, and after the COVID-19 pandemic, with an emphasis on RSV’s recent early onset severe seasons (2021/2022, 2022/2023 and 2023/2024), this study aims to elucidate epidemiology and transmission patterns across various age cohorts, to support targeted preventative programmes.^26^ ^28^ ^33^ Insights from this research are intended to inform public health authorities and health service planners to enhance timely preventive measures to alleviate RSV’s impact on the healthcare systems.^33^ Globally, a shift towards a *‘life-course approach to population health’* may address the burden of RSV in paediatric populations while mitigating outcomes in older adults, as peaks in paediatric cases frequently precede those in older adults.

Analyses of national-level and age-stratified baseline epidemiological data on RSV notification from the pre-immunisation decade would potentially serve as a baseline to compare the impact of immunisation approaches adopted on the island of Ireland. While an immunisation approach for newborn infants was adopted through the ‘Pathfinder Programme’ in the Republic of Ireland, a maternal vaccination strategy was adopted in the United Kingdom including Northern Ireland. As both commenced during the 2024/2025 season in two adjacent geographic regions sharing similar climatic and socioeconomic conditions, variations, if any, observed in RSV notifications and RSV-associated hospital admissions, based on two differing primary preventative strategies would be of public health relevance^. 20 23 24 34^

We hypothesised that RSV transmission follows a sequential pattern from infants and young children to older adults, with the objective of describing age-specific RSV notification trends over twelve seasons in Ireland, comparing pre- and post-COVID-19 patterns to inform national prevention strategies.

## METHODS

### Study design and data collection

This surveillance time-series study examines the national-level RSV notification trends in Ireland, comparing post-COVID-19 trends with pre-pandemic patterns. Comprehensive and prospectively collected weekly notification data on all laboratory-confirmed RSV notifications from week 40 in 2012 to week 20 in 2024 were obtained from the Health Protection Surveillance Centre (HPSC), Ireland’s authoritative national body on all hazards reporting and surveillance. Since RSV was made a statutory notifiable disease in Ireland in January 2012, the HPSC has systematically reported on all RSV cases.

### Variables analysed

Key variables included peak timing and magnitude, incidence rates per 100,000 population, and seasonal trends. These variables were stratified by age group (infants <1 year, children aged 1-4 years, and adults aged ≥65 years) to assess demographic-specific patterns. To ensure complete coverage, laboratory-confirmed RSV cases recorded in Ireland’s Computerised Infectious Disease Reporting (CIDR) were referred to as well.^24^

### Calculation of Incidence Rates

RSV incidence rates per 100,000 population were calculated using national census data from the Central Statistics Office (CSO), allowing for adjustments relative to population changes over time.^35^

### Data reporting and statistical analysis

Weekly data of laboratory-confirmed RSV notifications by age group were compiled to analyse trends and patterns, focusing on shifts between pre- and post-COVID-19 RSV seasons.

### Data protection and Research Ethics Approval

Data handling adhered to GDPR Article 9, ensuring lawful health data processing for surveillance.^36^ All data were securely stored following ISO 27001:2013, with data protection impact assessments (DPIAs) conducted to manage risks. Only deidentified and fully anonymised aggregate national-level surveillance data were analysed. The University of Limerick Hospital Group (ULHG) Research Ethics Committee (REC) waived the need for formal approval, based on the exemption clause for the deidentified notifiable disease data analysis approved by the Health Service Executive (HSE) in Ireland, founded on the Infectious Diseases Regulations of 1981 (amended in 2024 as number 528).

### Statistical analysis

Descriptive statistics and visualisations of onset and peaks of RSV infections by season compared pre-COVID-19 and post-COVID-19 trends. Poisson regression models were used to compare the annual incident rate (IR) per 100,000 population post-COVID-19 (2021-22, 2022-23 and 2023-24) to pre-COVID (2013-2019) for the age cohorts (<1 year, 1-4 years and ≥65 years). The Poisson models computed IRs with associated 95% confidence levels as well as incident rate ratios (IRR) to compare each year post-COVID-19 to average RSV incidence during 2013 – 2019.

Cross-correlation function (CCF) analysis was used to examine the strength of the association between weekly RSV incidence rates in older adults (≥65 years) and infections in the <1 year and 1-4 year age groups at different lag weeks. This approach identifies whether increases in one age group tend to precede or follow increases in another. For example, a high correlation at a lag of -2 weeks would indicate that the time series have similar weekly patterns, but one series leads the other by two weeks. In preparation for the CCF analysis, the weekly time series RSV rates per 100,000 in <1-year-olds, 1–4-year-olds and ≥65-year-olds were detrended to ensure the analysis focused on the cyclical component without interference from trends.

Statistical Analysis was undertaken using R (R Core Team 2020) and IBM SPSS V29 software. A 5% level of statistical significance was used throughout the analyses.

### Patient and Public Involvement

While direct patient and public involvement (PPI) was not applicable for analysing a nationally collected RSV notification and surveillance of a fully anonymised data, representation by the designated patient delegate of our Limerick Neonatal Charity (Irish Charity Registration No: 20073732) was ensured as an independent observer and offered parents’ views from the very outset of the design, drafting and conduct of the study.

### Reporting guidelines

We used the STROBE reporting guideline checklist during the manuscript editing.^37^

## RESULTS

### Overall RSV notifications in Ireland from 2012-2024

Analysis of RSV notification rates over time (Figure 1) reveals significant shifts in the temporal patterns and overall burden of RSV infections in Ireland from week 40 of 2012 to week 20 of 2024. Particularly in the post-COVID-19 era (up to and including the 2023/2024 season), overall RSV notifications have demonstrated a clear trend toward earlier peaks and heightened infection loads. In contrast to pre-COVID-19 pandemic years, when RSV peaks generally occurred in late winter (late December/early January), recent data from the 2021/2022-2023/2024 seasons, show a shift with peaks occurring as early as mid-November.

**Figure 1:**
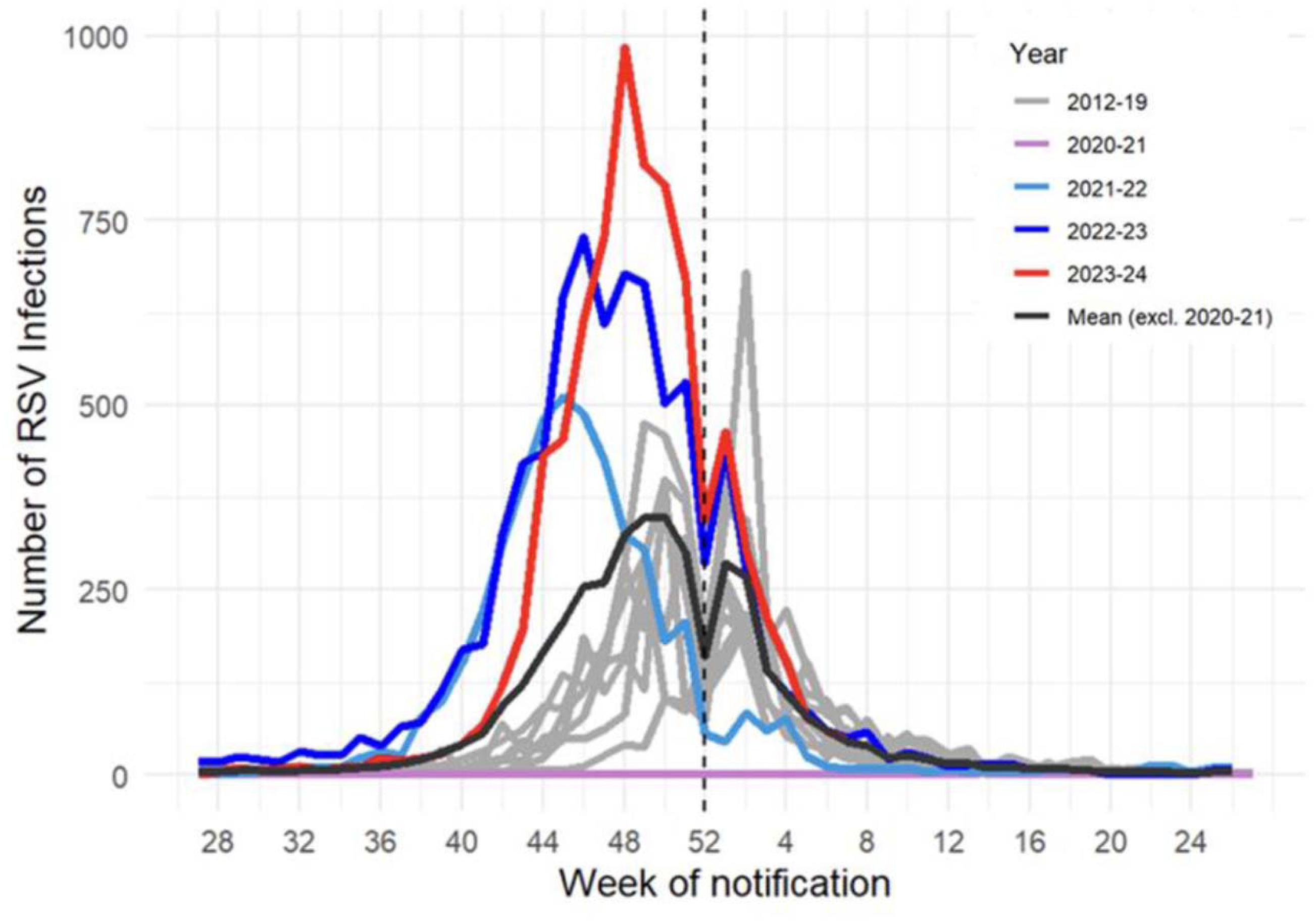
Total RSV notifications across study years (2012-2024)

The RSV epidemic peak for the 2022/2023 season occurred in Week 46 2022 (November 14th - November 20th) with 729 notified cases per week, while the 2023/2024 season saw an even higher peak of 985 notified cases per week, which occurred slightly later in Week 48 2023 (November 27th - December 3rd). This trend reflects a marked increase in incidence and an earlier seasonal onset post-COVID-19 pandemic for the 2021/2022, 2022/2023, and 2023/2024 seasons, with peaks occurring approximately 8 weeks after the initial rise in RSV cases.

Quantitatively, notified RSV cases increased significantly each year post-2020 2020 (up to week 20 2024). Compared to 2020-21 (a season with only a small number of sporadic RSV cases notified), the notification rate increased by 100% in 2021-22, with 510 cases per peak week (week 44). This was followed by a 1.43-fold increase in 2022-23 (729 cases in week 46), and a 1.93-fold increase in 2023-24 (985 cases in week 48). Additionally, peak incidence rates concentrated more intensely within a narrow window: RSV notifications within a three-week window surrounding the peak reached 96% in 2021-22, 77% in 2022-23, and 78% in 2023-24. Table 1 presents the annual incident rate (IR) per 100,000 (95% CI) for the age groups (<1 year, 1-4 years and ≥65 years) for each post-COVID-19 season (2021-22, 2022-23, and 2023-24) and incident rate ratios (95% CI) comparing the IR post-COVID-19 to the average IR pre-COVID.

**Table 1:** Peak weeks, cases, and annual incident rate analysis comparing RSV pre-and post-COVID-19. Annual Incident rates (95% CI) per 100,000 population from Poisson regression models. ^1^Incident rates computed using data up to Week 7 2024. ^2^Range (min-max) given for peak weeks and cases in 2013 – 2019.

|  | **Peak Week** | **Peak Cases** | **Incident Rate (95% CI)** | **Incident Rate ratio (95% CI)** |
| --- | --- | --- | --- | --- |
| **<1 year olds** |  |  |  |  |
| 2023/24<sup>1</sup> | 48 | 390 | 4993.43 (4814.53, 5178.97) | 1.87 (1.79, 1.94) |
| 2022/23 | 46 | 271 | 4289.22 (4123.66, 4461.44) | 1.60 (1.53, 1.67) |
| 2021/22 | 44 | 233 | 3324.93 (3184.73, 3471.29) | 1.24 (1.19, 1.30) |
| 2013 - 2019<sup>2</sup> | 49 - 1 | 200-300 | 2677.15 (2629.01, 2726.17) | *reference* |
| **1-4 year olds** |  |  |  |  |
| 2023/24<sup>1</sup> | 48 | 341 | 911.12 (873.54, 950.32) | 4.12 (3.91, 4.34) |
| 2022/23 | 46 | 207 | 697.33 (664.55, 731.73) | 3.15 (2.98, 3.34) |
| 2021/22 | 46 | 164 | 560.06 (532.49, 589.05) | 2.53 (2.39, 2.68) |
| 2013 - 2019<sup>2</sup> | 49 - 1 | 55 – 121 | 221.24 (214.63, 228.06) | *reference* |
| **≥65 years old** |  |  |  |  |
| 2023/24<sup>1</sup> | 51 | 160 | 174.03 (164.99, 183.56) | 3.96 (3.70, 4.25) |
| 2022/23 | 51 | 218 | 266.64 (255.4, 278.38) | 6.07 (5.71, 6.46) |
| 2021/22 | 49 | 43 | 61.8 (55.99, 68.21) | 1.41 (1.26, 1.57) |
| 2013 - 2019<sup>2</sup> | 2 – 4 | 6 - 83 | 43.92 (42.02, 45.91) | *reference* |

### RSV notifications in infants in Ireland 2012-2024

In infants aged <1 year, RSV infection peaks varied significantly across study years, highlighting post-pandemic shifts in infection timing and intensity (up to and including the 2023/2024 season) (Figure 2). During the 2019–2020 season, RSV notified cases peaked in the second week of 2020, reaching 305 cases per week, before declining. No RSV peaks were observed during the 2020–2021 season, coinciding with COVID-19 restrictions. In the 2021–2022 season, RSV notifications peaked earlier in Week 44 (November 1st–7th), with 233 cases per week. Subsequent seasons showed further increases in both peak timing and intensity, with 271 cases per week in 2022–2023 (Week 46, November 14th–20th) and a notable peak of 390 cases per week in 2023–2024 (Week 48, November 27th–December 3rd).

**Figure 2:**
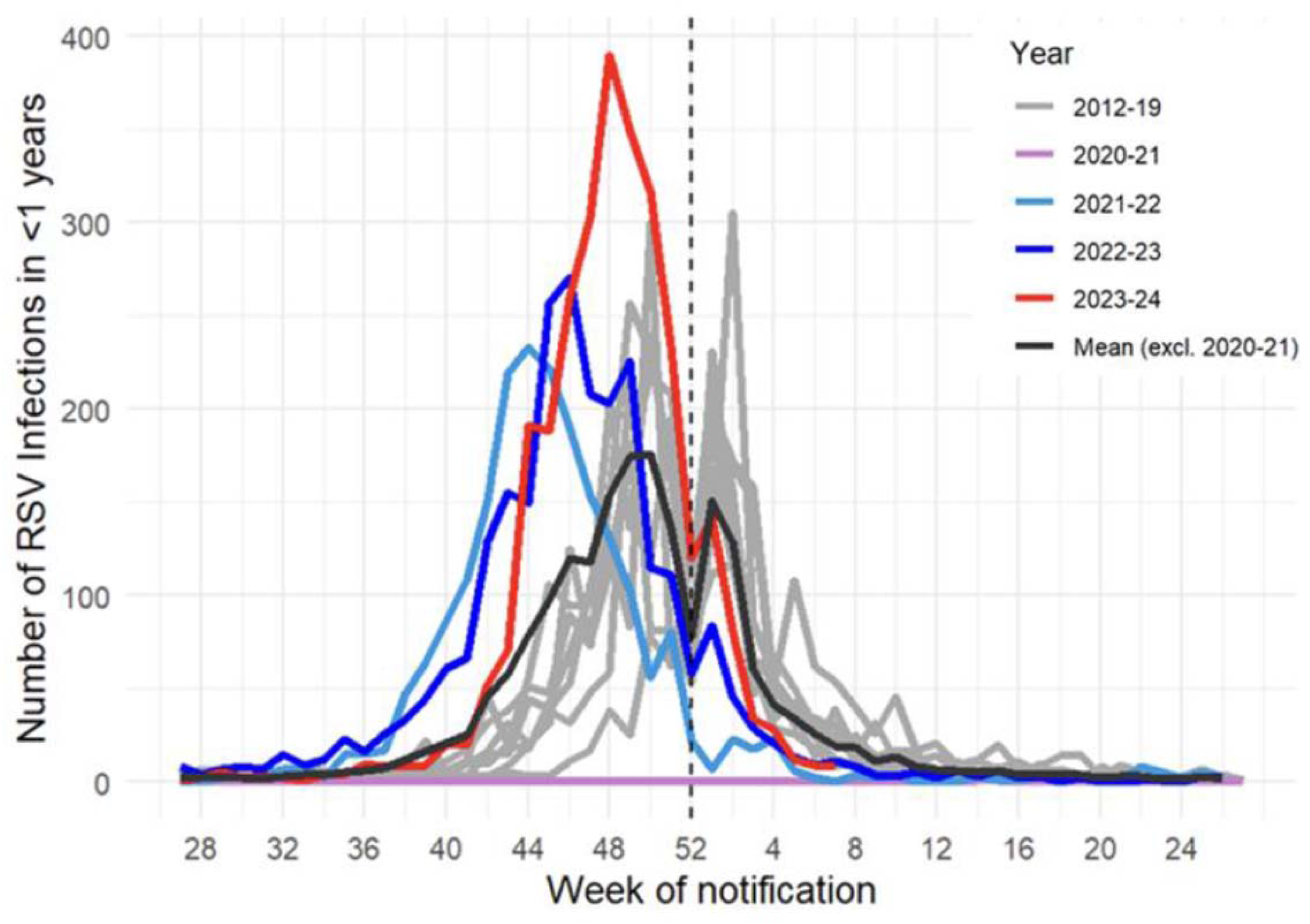
RSV notifications in infants (aged<1 year) across study years (2012-2024)

Compared with the low infection rates of 2020–2021, RSV cases increased to 3324.93 per 100,000 (95% CI: 3184.73, 3471.29) in 2021–2022 almost 25% higher than pre-COVID-19 years (IRR = 1.24, 95% CI: 1.19, 1.30). Incidence levels increased further in 2022–2023 and again in 2023–2024 (IR = 4993.43 (4814.53, 5178.97). Additionally, concentrated peaks were evident within three weeks surrounding each season’s highest infection rate. Specifically, the percentage of annual RSV cases within this peak window was 84% (478/566) in 2021–2022, 88% (561/637) in 2022–2023, and 84% (751/899) in 2023–2024, showing a compressed period of high transmission. These findings indicate a marked shift towards higher, earlier, more concentrated and apparently shorter-tailed RSV infection peaks in infants <1-year in post-pandemic seasons (up to and including the 2023/2024 season), underscoring the need for vigilant RSV monitoring and early intervention strategies in this vulnerable age group. Similarly, increasing levels of RSV were recorded in the 1-4 year-old group post-COVID-19 (Figure 3 and Table 1), with 2023-34 IR = 4993.43 (4814.53, 5178.97), over 4 times higher than pre-COVID-19 incidence levels (IRR = 4.12, 95% CI: 3.91, 4.34).

**Figure 3:**
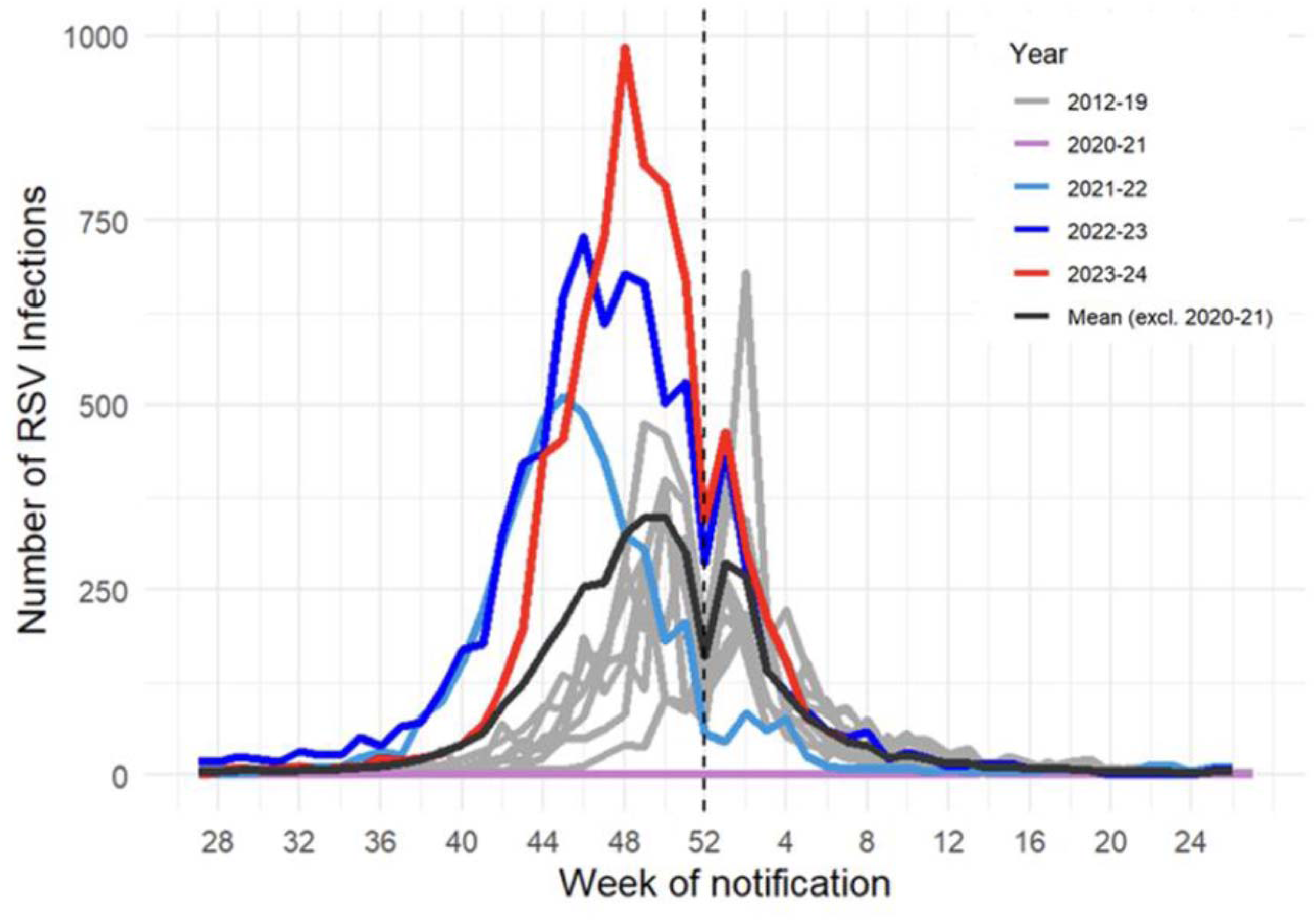
RSV notifications in children aged 1-4 years for 2012-2024

### RSV notifications in Ireland for ≥65 years

For adults aged ≥65 years, RSV peak timing and infection intensity also shifted during the post-pandemic (2021/2022 - 2023/2024) seasons (Figure 3). Peaks in notified cases rose significantly, with a 5.07-fold increase in 2022-23 compared to 2021-22, with incident rates 6 times higher in 2022-23 compared to pre-COVID-19 (IRR = 6.07; 95% CI: 5.71, 6.46). Seasonal peaks for this age group typically occurred four weeks after peaks in notified cases in young children, (Table 1 and Figure 4), highlighting a distinct age-specific transmission lag.

**Figure 4:**
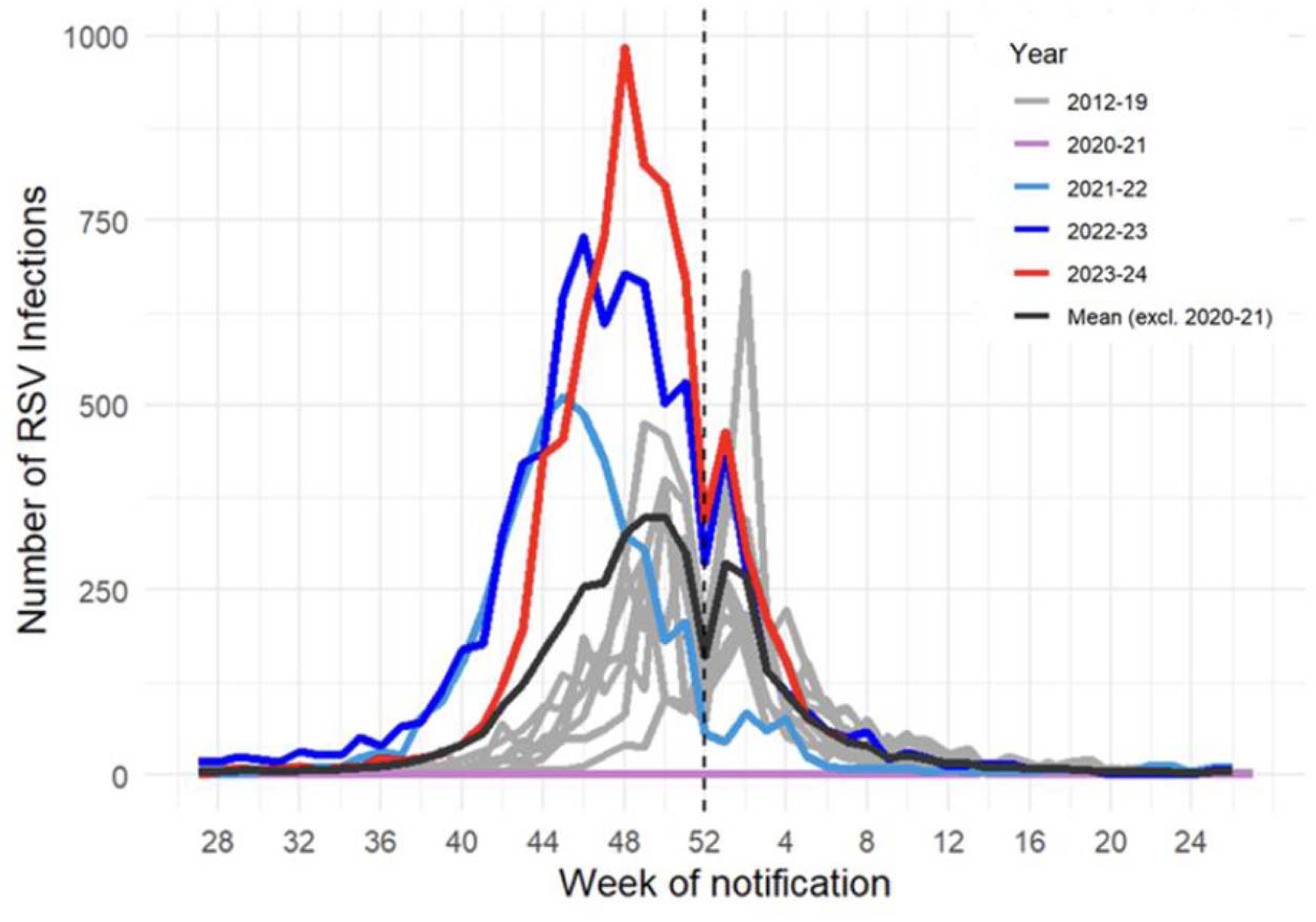
The number of RSV notifications in Ireland for older adults ≥65 years of age.

Key observations suggest an intensified, age-related transmission pattern in RSV post-COVID-19, with implications for targeted RSV prevention and timely interventions.

### Cross-correlation function (CCF) analysis of time lags between RSV rates in children aged <1 year, 1-4 years, and older adults ≥65 years

The time-lag analysis of detrended RSV infection rates in ≥65 years vs <1 year and vs 1-4 years from 2012 to 2024 found infections in ≥65 years were strongly correlated to infections in the younger age groups lags of between 3 and 5 weeks. The CCF plots, Figure 5, show the highest correlation to occur at a week lag 4 for both <1 year olds (r = 0.79) and 1-4 year olds (r = 0.86). This analysis captures the concept of an ‘*RSV march’*, where infections initially rise in infants and younger children and subsequently spread to older adults after a few weeks.

**Figure 5:**
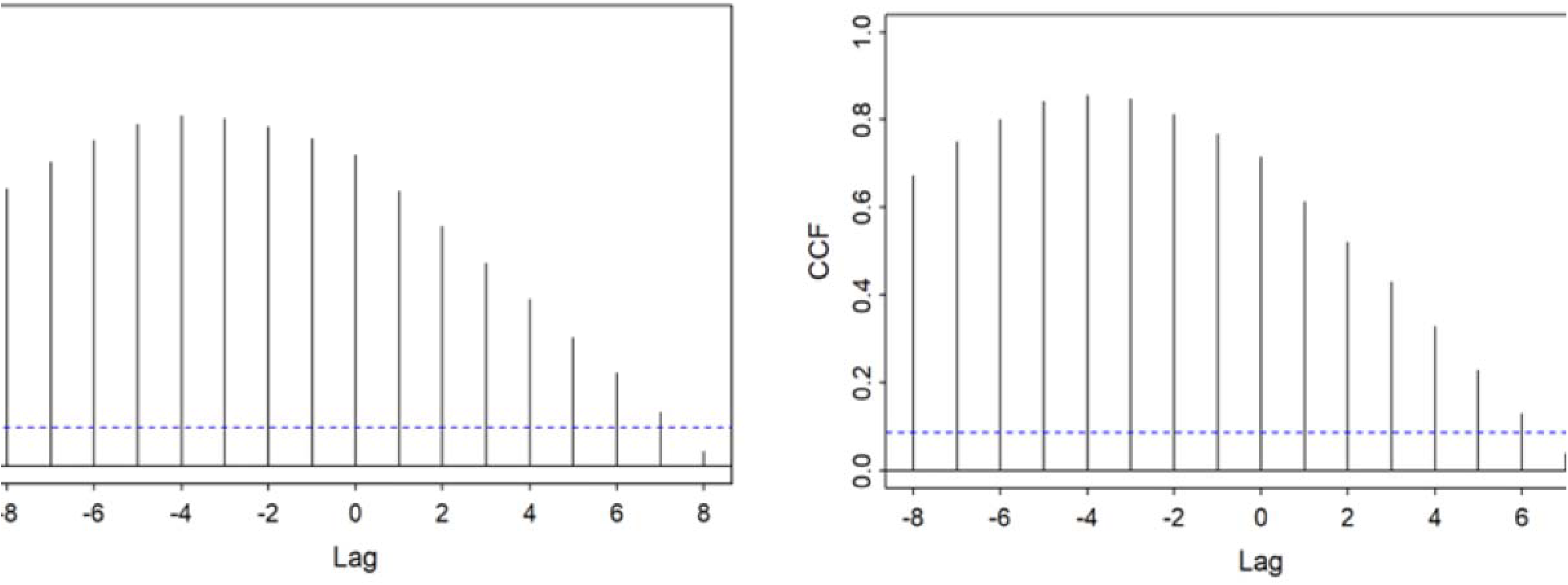
Time-lag analysis of RSV notifications 2012 – 2024 (excluding 2020-21) in ≥65 years old vs <1-year-olds (Figure 5a) and vs 1-4-year-olds (Figure 5b).

## DISCUSSION

In this study, we found a clear shift toward earlier and more intense RSV peaks in the post-COVID-19 period, particularly among infants (<1 year) and younger children (1–4 years), with a consistent 3–5 week lag indicating that peaks in these age groups preceded those in older adults (≥65 years).

### Graphical Insights from National RSV Surveillance Data

National RSV surveillance data provides compelling insights into RSV incidence across age groups, highlighting the elevated vulnerability of infants under one year of age. Post-COVID-19 pandemic trends for the 2021/2022, 2022/2023 and 2023/2024 seasons reveal a shift in RSV seasonal timing, with infections peaking earlier in November and December—approximately eight weeks after initial rises in case numbers. The rise in incidence rates is particularly evident among children under five years, whose incidence rates in 2022 and 2023 surpassed pre-COVID-19 pandemic levels. This shift aligns with broader European observations up to and including the 2023/2024 season, where atypical and earlier RSV seasons post-pandemic were noted across countries, while pre-pandemic RSV seasonality patterns were described in earlier European, North American and Global studies,^38^ ^39^ showing intensified and altered RSV patterns due to shifts in social behaviours and public health measures. In adults >65 years, RSV incidence peaks approximately four weeks after the peak in infants and younger children, highlighting the importance of age-stratified timing in healthcare planning and resource allocation (Figure 4^).30^

These observed shifts in RSV incidence across age groups highlight the need for age-targeted preventive strategies during RSV-endemic seasons, particularly for high-risk groups such as infants. As shown in this study, infants and young children are predominantly driving the initial wave of RSV transmission each winter, with the incidence in older adults who are also at risk occurring afterwards. The increased RSV burden among infants supports immunisation of newborn infants with long-acting mAb or maternal RSV vaccination as primary preventive measures. Initiatives such as breastfeeding promotion and avoidance of cigarette smoke exposure should also be considered; together, these interventions act as adjuncts to immunisation programmes, assisting to mitigate the RSV-related healthcare demands evident in surveillance data, reducing the hospital admissions as well as pressure on the scarce PHDU and PICU beds during peak RSV epidemic periods. These measures may also interrupt transmission chains thereby reducing incidence in older adults, and mitigating the RSV-related healthcare burden in older adults.

The sequential progression of RSV transmission across age groups illustrates how RSV activity in younger children, particularly infants and those aged 1-4 years, frequently precedes increases in cases among older adults (≥65 years) by a lag of 3-5 weeks (Figure 5). Cross-correlation analysis, with high correlation coefficients (0.81-0.82 for children aged 1-4 years and 0.72-0.75 for infants aged <1 year), further substantiates this transmission pattern, indicating that early RSV peaks in paediatric populations often signal subsequent waves in older adults, with similar parallels found across Europe. ^38^ ^39^ This ‘*RSV march’* or transmission pattern across age groups, positions younger children as the primary source of RSV transmission cycles, and preventive strategies targeting this age group can effectively reduce household and community transmission rates with a resultant proportional reduction in hospital admissions. By taking advantage of this predictable progression and interrupting the RSV transmission cycle, through preventative measures such as immunisation, both infants and older adult populations could be protected without necessitating resource-intensive interventions. Immunisation strategies such as neonatal long-acting monoclonal antibodies or maternal RSV vaccination may have the potential to significantly decrease RSV transmission from infants to other household members, particularly older adults vulnerable to severe RSV outcomes, underpinning the ‘herd immunity’ or ‘community immunity’ principles.^40^ ^41^ A targeted approach, which focuses resources on the population initiating transmission cycles each season, may yield widespread protective benefits across demographics while optimising the allocation of healthcare resources.

### ‘Infant to older adult RSV march’ as a predictive tool and new immunoprophylaxis initiatives

Cross-correlation analysis shows that RSV infections in children, particularly those aged less than five years, often precede increases in older adults by a consistent 3-5 week lag. This predictive window provides a critical opportunity to anticipate and prepare for surges in RSV-related care needs, enabling healthcare systems to optimise resources during peak RSV season.

Following the severe 2022/2023 and 2023/2024 RSV seasons, Ireland introduced the ‘RSV immunisation Pathfinder *programme’* for newborn infants, born between September 1, 2024, and February 28, 2025. This initiative aims to reduce RSV-related hospitalisations and critical care needs among infants, who are at the most risk of severe disease. Similarly, the UK launched a maternal immunisation programme against RSV in September 2024, conferring protection to infants through maternal antibodies, while nirsevimab remained targeted to specific high-risk infant groups, such as those born <32 weeks’ gestation. In addition, the UK also implemented an RSV vaccination programme for older adults in September 2024.^42^

These RSV immunisation programmes commencing during the 2024/2025 season, address RSV transmission at different stages and ages, and should potentially reduce the overall burden of RSV. Immunisation with nirsevimab in newborns should potentially prevent early RSV peaks in infants, reducing RSV hospitalisations and ICU admissions and may potentially reduce overall transmission of RSV in the community for other older age groups.

By leveraging the predictive implications of RSV epidemic waves, health systems in Ireland and the UK are poised to optimise resource allocation, enhance critical care preparedness, and support coordinated, age-specific interventions. This forward-looking approach could play a pivotal role in mitigating the seasonal impact of RSV on healthcare systems. The two varied approaches adopted in Ireland and UK also offer a ‘real world’ opportunity to analyse the proportion of the target population who opted for the protective strategy, i.e. number of newborn infants who received nirsevimab among the total eligible births during the ‘*Pathfinder programme’* in Ireland vs the number of pregnant women >28 weeks who opted to get the maternal vaccination among the total eligible pregnant population from 1^st^ September 2024 in the UK.^43^

Ireland’s approach to immunise newborn infants against RSV merits weighing against the UK’s strategy of maternal vaccination, for which the historic rates across the globe remain suboptimal.^44^ ^45^ ^46^ For influenza, maternal vaccine uptake is only around 50%^45^ with hesitancies being attributed to a myriad of factors including socioeconomic status, public health information, and ethnic and racial disparities.^45^ ^47^ Additionally, maternal vaccinations have been shown to require a significantly higher uptake to achieve comparable reductions in disease burden achieved through direct immunoprophylaxis offered for all infants.^44^ ^45^ ^47^ Spain was one of the first EU countries to administer nirsevimab to infants and their coverage reached over 90%, suggesting realistic expectations for broad implementation.^46^ Infant administration of nirsevimab in Germany compared to maternal vaccination reported that nirsevimab in infants provides broader coverage by immunising infants born before and during the RSV season.^45^ Specifically, targeting infants aged 1–5 months, as opposed to those older than 6 months, reduced infection rates by an additional 18%. In contrast, maternal vaccines cannot passively immunise infants already born well before the RSV season, resulting in a narrower reach for maternal vaccination at comparable uptake levels.^45^ This difference highlights the plausible positive benefits of seasonal long-acting monoclonal antibody strategies, such as nirsevimab or clesrovimab immunoprophylaxis, in reaching a larger proportion of the target population and preventing substantially more RSV-associated hospital admissions. Moreover, the decline in RSV infections among newborn infants achieved through either strategy may also indirectly reduce transmission to older adults.

Post-hoc analysis following the 2023-2024 winter season nirsevimab immunisation of newborn infants yielded >90% uptake and resultant >70% reduction of RSV-associated hospitalisation of infants.^41^ ^48^ ^49^. Initial trends in Ireland suggest a similar uptake, at one of the six designated health regions of Ireland (University of Limerick Hospital Group), of the 766 eligible newborn infants from 1^st^ September to 20^th^ November, 690 received nirsevimab (90%). Early reports from HPSC and ECDC are encouraging regarding the surveillance data for weeks 35-45 of 2024 having reduced numbers compared to the corresponding period of 2023.^50^ ^51^ Whether this reflects an impact by the ‘*Pathfinder programme’* or a delayed start of the season or the natural normalisation of the peak incidence to the pre-COVID-19 era (‘*realignment of immunity gap’*), all remain possibilities for the time being.

### Immunity Gap

Epidemiology of RSV in the post-COVID-19 period demonstrates the concept of an ‘immunity gap’. Historically most children had an RSV infection by year two, spreading to others and maintaining viral circulation and resultant immunity. However, those infants born during COVID-19 remained *‘RSV naïve’* due to the COVID-19 mitigation measures, remaining vulnerable for RSV infection during early childhood. This shift is reflected in recent trends across multiple regions, where RSV activity has increased in both timing and intensity following the pandemic.^5^ Similarly, a Danish national study up to January 2023 observed that older children were more likely to be admitted during the 2021-2022 ^57^ and 2022-2023 seasons, and a large US study noted that children admitted for bronchiolitis during these seasons were older, more likely to require ICU admission, and more likely to receive non-invasive ventilation.^5^ ^58^ ^59^ While this shift in age distribution may result in reduced severity among older children experiencing RSV for the first time, infants who missed the opportunity to acquire maternal antibodies due to fewer maternal exposures during the pandemic are now more vulnerable to severe RSV disease.^49^ ^60^ This emphasises the need for targeted interventions to protect high-risk groups, particularly infants unprotected by maternal antibodies.^61^ ^62^ This also emphasises the need for further strengthening of RSV surveillance systems in Ireland, to ensure that any potential shift in RSV epidemiology because of immunisation programmes is actively monitored.

### Climate

While some studies have linked RSV seasonality to climatic factors, a single, definitive factor that explains these periodic patterns remains unidentified.^14^ ^15^ Low temperature and high humidity are key predictors of RSV epidemics.^13^ ^14^ Nevertheless, while Ireland in the last 5 years has noted increased warming and precipitation, mirroring global climate change, there has been a significant shift during the 2022/2023 and 2023/2024 seasons where RSV commenced its activity earlier and reaching its peak earlier than previously observed.^14^ Research indicates a significant correlation between high annual precipitation and RSV peak activity, suggesting that a deeper understanding of RSV seasonality could optimise the timing of intervention strategies.^26^ ^27^ For example, in Northern Brazil, annual vaccination campaigns are scheduled according to the Southern Hemisphere winter, but this timing is too late for the region’s local influenza season.^27^ Thus, climate and environmental conditions are also essential considerations for future RSV immunisation programmes.

### Limitations

This study has several limitations that should be noted. Firstly, while the data analysed was part of a national RSV notification and surveillance programme in Ireland, the reliance on a retrospective data analysis covering over a decade may introduce biases and inaccuracies due to potential inherent inconsistencies in data quality and completeness. Additionally, by focusing on the incidence of RSV notifications within specific age groups, this study did not capture other age groups (i.e. those aged 5-64 years), or factors such as underlying health conditions, and socioeconomic or geographic factors that could affect incidence rates.

Moreover, the seasonal patterns observed in this study may be influenced by external variables not accounted for, such as increased use of multiplex PCR, changes to laboratory testing capacity, improvements to surveillance and reporting (e.g. use of robotic process automation for RSV notifications), changes to healthcare-seeking and testing behaviour and climate variability. Several studies in the literature indicate that climate has a significant effect on RSV peaks, so much so that some countries observe biannual RSV epidemics versus annually.^26^ ^27^ ^28^ Thus, the trends observed in our nationwide study cannot be applied to all European countries, as diverse environmental factors can affect viral activity at even small geographical scales.^27^ While these associations have not been firmly established, further investigation on the effects of environmental factors is warranted.^26^

This analysis is limited to laboratory-confirmed RSV notifications only, and analyses of GP visits and ED presentations were not included, which may introduce selection bias. Such bias would likely bias estimates toward the null, underestimating the true incidence of RSV in the community, though the magnitude of this bias cannot be quantified from available data. Data on hospital admissions, length of stay (LOS), or critical care admissions/bed utilisation in PHDU or PICU were not available. This study also did not include epidemiological or demographic characteristics of notified cases including sex, comorbidities, rurality, socioeconomic status, premorbid health status, breastfeeding rates, and prematurity, to better understand RSV epidemiological changes pre and post-COVID-19 pandemic^5^ and to assess their impact on RSV trends. Sex may act as a risk modifier, and sex-related differences in RSV severity warrant further study. Although we aimed to predict the timing of future RSV surges, inherent uncertainties in infectious disease forecasting may affect the accuracy of these projections.

A key limitation in RSV research is the difficulty of comparing pre- and post-pandemic (COVID-19) surveillance data due to changes in testing practices.^19^ Before the pandemic, RSV testing was less commonly used in acute hospital settings, as treatment is generally supportive and does not always require specific testing.^19^ However, the COVID-19 pandemic has led to wider availability and use of multiplex testing, making post-pandemic RSV data more robust and frequent than in previous years.^19^

## CONCLUSION

This study offers important insights into the incidence and seasonality of RSV across age groups over the past decade in Ireland, with a particular focus on infants (<1 year), young children (1–4 years), and older adults (≥65 years). By analysing RSV trends before and after the COVID-19 pandemic, we have observed how public health measures and changes in social behaviour altered the transmission dynamics, revealing a sequential spread from infants and young children to older populations. These findings may inform and serve as baseline data in the evaluation of the RSV immunisation ‘*Pathfinder programme’* of 2024/2025 in Ireland, by monitoring the impact of this primary prevention in reducing RSV notifications and hospitalisations.

The provision of timely RSV surveillance and immunisation uptake data, across the life spectrum, to policymakers and public health authorities is essential for informing the RSV immunisation policies. This is best achieved through a robust implementation strategy using a *life-course approach to public health*, and an effective awareness campaign to target populations. Further analysis of RSV-associated ALRI and hospital admissions avoided among infants at the end of the 2024/2025 RSV season will inform and potentially guide the most appropriate immunisation approach for Ireland. International studies and comparisons with countries using alternative RSV preventative strategies (newborn immunoprophylaxis / maternal vaccination/vaccination of older adults) will also inform future immunisation allocations in Ireland.

## Acknowledgements

Authors wish to acknowledge the support form Dr Joan O’Donnell and staff at the Health Protection Surveillance Centre (HPSC), as well as the Public Health Department at the Health Service Executive (HSE) for clarifying information on the ‘*Pathfinder programme’* for RSV immunisation of newborn infants in Ireland. The authors acknowledge the editorial support offered by Dr Gwinyai Masukume in the preparation of the final manuscript.

